# Disentangling the relationship between biological age and frailty in community-dwelling older Mexican adults

**DOI:** 10.1101/2024.08.20.24312308

**Authors:** Carlos A. Fermín-Martínez, Daniel Ramírez-García, Neftali Eduardo Antonio-Villa, Miriam Teresa López-Teros, Jacqueline A. Seiglie, Roberto Carlos Castrejón Pérez, Carmen García Peña, Luis Miguel Gutiérrez-Robledo, Omar Yaxmehen Bello-Chavolla

## Abstract

**OBJECTIVE:** Older adults have heterogeneous aging rates. Here, we explored the impact of biological age (BA) and accelerated aging on frailty in community-dwelling older adults.

**METHODS:** We assessed 735 community-dwelling older adults from the Coyocan Cohort. BA was measured using AnthropoAge, accelerated aging with AnthropoAgeAccel, and frailty using both Fried’s phenotype and the frailty index. We explored the association of BA and accelerated aging (AnthropoAgeAccel ≥0) with frailty at baseline and characterized the impact of both on body composition and physical function. We also explored accelerated aging as a risk factor for frailty progression after 3-years of follow-up.

**RESULTS:** Older adults with accelerated aging have higher frailty prevalence and indices, lower handgrip strength and gait speed. AnthropoAgeAccel was associated with higher frailty indices (β=0.0053, 95%CI 0.0027-0.0079), and increased odds of frailty at baseline (OR 1.16, 95%CI 1.09-1.25). We observed a sexual dimorphism in body composition and physical function linked to accelerated aging in non-frail participants; however, this dimorphism was absent in pre-frail/frail participants. Accelerated aging at baseline was associated with higher risk of frailty progression over time (OR 1.74, 95%CI 1.11-2.75).

**CONCLUSIONS:** Despite being intertwined, biological accelerated aging is largely independent of frailty in community-dwelling older adults.

## INTRODUCTION

Despite recent advances in the understanding of human aging across the lifespan, its study in older adults remains challenging^1,2^. Differences in the rates of biological aging in older adults with similar chronological age (CA) have been characterized using methodologies that capture age-related changes in physical function, independence, and resilience with greater nuance compared to CA^3^. Specifically, older adults present heterogeneous aging profiles, ranging from modest physiological changes to significant impairments in physical function, disability and dependence; entities that may be influenced by underlying comorbidities and biological mechanisms independent of CA^4–6^. Amongst available approaches to capture aging are the concepts of biological age (BA) and frailty, which are often treated interchangeably and have been proposed to share common pathways^7,8^.

Recent data has shown that BA changes in otherwise healthy midlife adults are associated with progressive accumulation of deficits, multimorbidity and frailty^9,10^. Data on the influence of BA on frailty phenotypes in older adults are scarce; however, evidence in centenarians suggests that such markers are useful to model aging even at extreme ages^11,12^. Recently, we developed AnthropoAge as a proxy of BA that captures multimorbidity and body composition changes associated with risk of 10-year mortality^13^. However, the application of AnthropoAge in older adults has not been reported, nor its association with frailty phenotypes. Here, we analyzed data from community-dwelling older adults in Mexico City to validate the use of AnthropoAge in older adults, aiming to characterize: 1) The extent to which accelerated biological aging intersects with frailty, 2) the impact of frailty on the body composition and functional phenotype of accelerated aging in older adults, and 3) the relevance of accelerated aging as a risk factor for progression to frailty.

## METHODS

### Study design and participants

We analyzed data from participants enrolled in the Coyoacán Cohort Study, a prospective population-based cohort of randomly selected community-dwelling adults ≥70 years from the municipality of Coyoacán in Mexico City. Complete study details are published elsewhere^14^. Briefly, baseline data collection was conducted from March 2008 until July 2009 in a two-stage process: first, participants were interviewed for sociodemographic and health-related information using standardized questionnaires, followed by medical and anthropometric examinations conducted by trained health personnel. A follow-up evaluation was conducted in 2011, whereby questionnaires were repeated, and vital status was ascertained by verbal autopsy of proxy relatives. Data collection protocols and study procedures were approved by the Ethics Committee of the Instituto Nacional de Ciencias Médicas y Nutrición Salvador Zubirán. For this sub-analysis, we included participants with complete anthropometric measurements and data to evaluate two frailty measures.

### Anthropometry and physical function measures

Standing height and weight were measured using Seca-214 stadiometers and Seca-803 scales. Mid-upper-arm, waist, hip, and calf circumferences were measured in centimeters using non-stretch fiberglass measuring tape on the left side of the body with participants standing. Maximal voluntary handgrip strength of the non-dominant hand was measured in kilograms with participants standing using a Baseline™ Smedley spring-type hand dynamometer. Body-mass index (BMI) was obtained by dividing weight in kilograms by the square of height in meters, the waist-to-height ratio (WHtR) by dividing waist by height in centimeters and the waist-to-hip ratio (WHR) by dividing waist by hip circumference in centimeters. Physical performance was assessed with gait speed calculated from the 4 m walk included in the Short Physical Performance Battery test^15^, and with the Timed Up & Go test, which measures the time in seconds as the participant rises from a chair without support, walks 3 meters, turns, walks back, and sits down again^16^. Grip strength, gait speed and Timed Up & Go data were only available for a subsample of n=283, n=267, and n=232 participants who underwent full medical evaluation, respectively. Estimates of all anthropometry measures are the average of at least two non-consecutive measurements^14^.

### AnthropoAge estimation as a proxy of biological age

AnthropoAge uses CA and anthropometric measures to predict sex-specific 10-year mortality risk as a proxy of BA^13^. For this analysis, we implemented the simplified version of AnthropoAge using the *AnthropoAgeR* package^17^, which uses CA in years, BMI, and WHtR at baseline. To estimate BA acceleration, we calculated AnthropoAgeAccel using residuals from a linear model regressing AnthropoAge onto CA^13,18^. Accelerated aging was defined as AnthropoAgeAccel ≥0 years.

### Frailty measures

Frailty is a multidimensional phenomenon, which encompasses both deficit accumulation and phenotypic changes. Because of its complexity, frailty has been operationalized using different approaches^19^. To increase generalizability of our findings, we implemented two distinct frailty measures:

1. **Modified frailty phenotype:** We used a modified definition of the frailty phenotype proposed by Fried et al. previously validated for this population^20,21^, which uses data from interview questionnaires to identify: a) Unintentional weight loss ≥5□kg in the last 12□months, b) Exhaustion, c) Low physical activity, d) Slowness, and e) Weakness. As previously reported, participants were categorized as frail if they fulfilled ≥3 criteria, pre-frail with 1–2 criteria, and non-frail with no criteria^22^.
2. **Frailty index:** We calculated the frailty index using Searle’s procedure which considers data from 42 deficits covering symptoms, signs, disabilities, and diseases^23–25^. Deficits considered are coded as binary variables and include: breathing difficulties, myocardial infarction, stroke, hypertension, cancer, diabetes, dyslipidemia, thyroid disease, fractures, arthritis, urinary incontinence, eyesight difficulties, hearing difficulties, falls, pain, smoking, difficulties from: pushing heavy objects, lifting a coin, being seated, standing up from a chair, preparing a meal, bathing, dressing, toileting, getting in and out from bed, moving around the house, eating, shopping, medication intake and making finances; restless sleep, happiness, loneliness, sadness, low energy, depressed, feeling everything is an effort, self-rated health, self-rated health compared a year ago and recent hospitalization. All deficits are then added and divided by the overall number of deficits resulting in a quotient which follows a gamma distribution, ranging from zero to one, with higher values representing higher frailty severity^25^.

### Statistical analyses

Categorical variables are reported as counts and frequencies, and continuous variables are reported as medians with interquartile ranges. Comparisons across categorical variables were conducted using Chi-squared or Fisher’s hypergeometric tests, while Wilcoxon signed rank tests were conducted for continuous variables. All analyses were conducted using R version 4.3.3 and a p-value<0.05 defined statistical significance.

### Association between frailty scores and AnthropoAge

To explore the association between the frailty index and the number of frailty components in the frailty phenotype (coded as 0, 1, 2 or ≥3) with AnthropoAge and AnthropoAgeAccel at baseline, we used the Spearman correlation coefficient (ρ). We explored the association of frailty indices with AnthropoAgeAccel at baseline using multivariable linear regression, adjusted for CA, sex, and number of comorbidities; additionally, the association between AnthropoAgeAccel and frailty categories (non-frail, pre-frail, and frail) was evaluated using ordinal logistic regression to estimate odds ratios (OR) of having more severe frailty categories, adjusted for CA, sex, and number of comorbidities at baseline. Next, we compared available anthropometric and physical performance measures in participants with and without accelerated aging (AnthropoAgeAccel values ≥0 vs. < 0) stratified by sex to investigate the influence of non-frailty versus pre-frailty/frailty on aging phenotypes^13^.

### Accelerated aging as a risk factor for frailty progression

We explored transitions across frailty phenotypes at baseline until the 3-year follow-up stratified by the presence of accelerated aging. To explore the influence of accelerated aging at baseline with these transitions, we fitted a mixed effects ordinal logistic regression using the *ordinal* R package to estimate ORs for transitions across frailty categories over time. Models were adjusted for sex, CA, and number of comorbidities at baseline.

## RESULTS

### Study population

From 1,124 participants recruited at baseline, we included 735 participants with complete anthropometric and frailty assessments. Amongst them, 389 (53%) were women, the median CA was 76 years (IQR 73-81), and 335 participants had accelerated aging (45.6%). Compared to those without, participants with accelerated aging had similar CA, but higher prevalence of frailty, higher frailty index, BMI and WHtR, and higher prevalence of diabetes (**Table 1**). Participants with accelerated aging also had lower handgrip strength and gait speed. We observed a strong correlation between AnthropoAge and CA (ρ=0.93, 95%CI 0.92, 0.94), without significant differences by sex (p=0.515, **Figure 1**). After follow-up, status of 586/735 (79.7%) participants were known, amongst whom 61 had died (10%) without differences in those with accelerated aging (**Table 1**).

**Figure 1.**
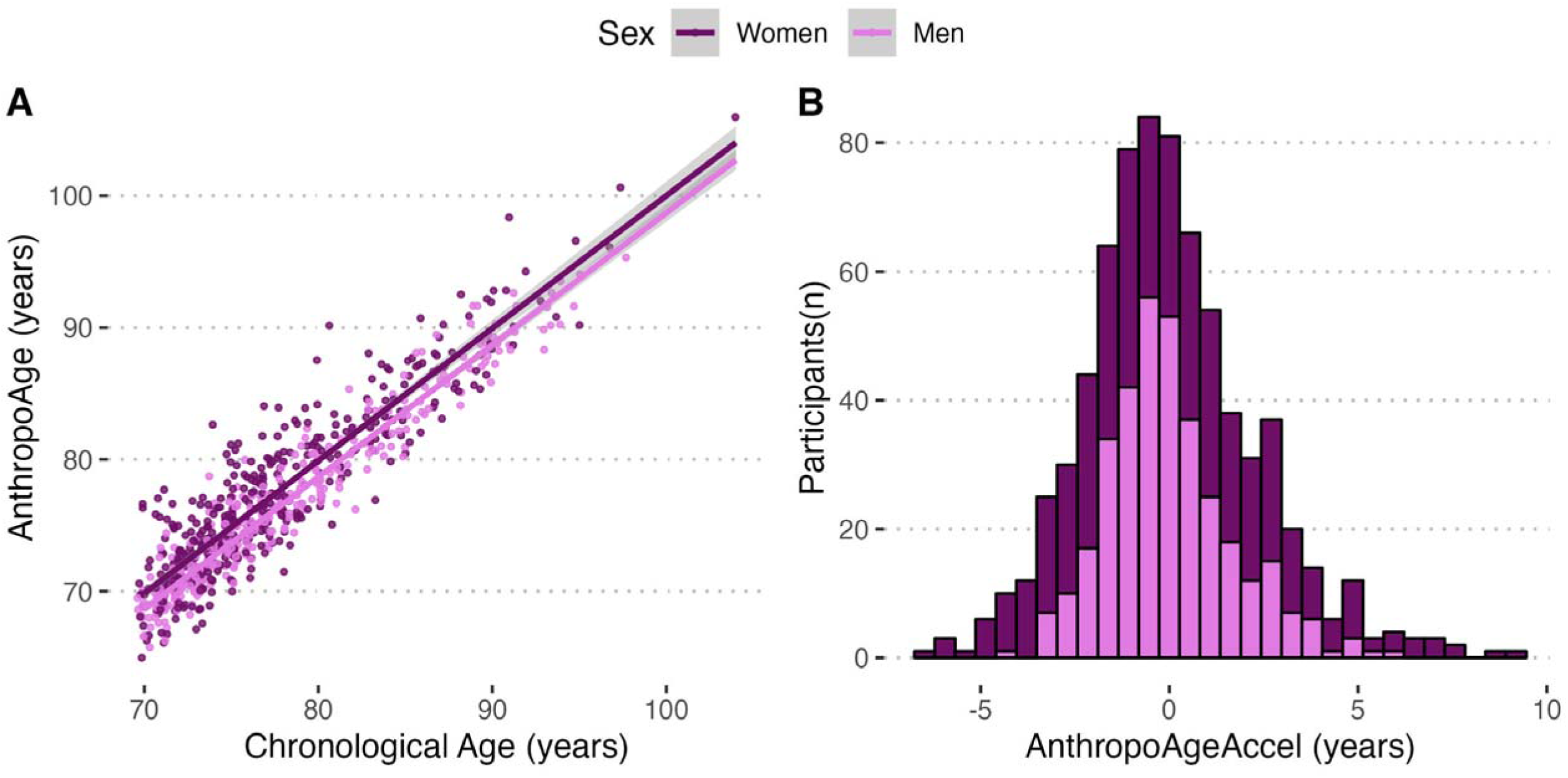
Relationship between Chronological Age (CA) and AnthropoAge values at baseline stratified by sex in 725 community-dwelling older adults from the Coyoacán Cohort (**A**). The figure also shows a histogram depicting the distribution of AnthropoAgeAccel values stratified by sex (**B**).

**TABLE 1.** Demographic and clinical characteristics of the study sample, categorized by the presence of accelerated aging as defined by AnthropoAgeAccel values 0 vs. ≥0 years.

| Characteristic | Overall Sample |  |  | p-value<sup>2</sup> |
| --- | --- | --- | --- | --- |
|  | Overall<br>N = 735<sup>1</sup> | Non-Accelerated<br>N = 400<sup>1</sup> | Accelerated<br>N = 335<sup>1</sup> |  |
| **Female sex** | 389 (53%) | 206 (52%) | 183 (55%) | 0.4 |
| **Chronological Age (years)** | 76.0 (73.0, 81.0) | 76.0 (73.0, 81.0) | 76.0 (73.0, 81.0) | 0.3 |
| **AnthroAge (years)** | 76 (72, 81) | 74 (71, 79) | 77 (74, 83) | <0.001 |
| **Frailty status** |  |  |  | <0.001 |
| Non-fragile | 366 (50%) | 221 (55%) | 145 (43%) |  |
| Pre-frail | 284 (39%) | 151 (38%) | 133 (40%) |  |
| Frail | 85 (12%) | 28 (7.0%) | 57 (17%) |  |
| **Frailty index** | 0.28 (0.23, 0.33) | 0.25 (0.23, 0.33) | 0.28 (0.23, 0.35) | 0.005 |
| **BMI (kg/m<sup>2</sup>)** | 26.8 (24.0, 29.5) | 26.4 (24.3, 28.4) | 27.3 (23.5, 31.2) | 0.034 |
| **Waist-to-height ratio** | 0.61 (0.56, 0.66) | 0.58 (0.54, 0.62) | 0.66 (0.60, 0.70) | <0.001 |
| **Waist-to-hip ratio** | 0.96 (0.90, 1.01) | 0.92 (0.87, 0.97) | 0.99 (0.95, 1.04) | <0.001 |
| **Calf circumference (cm)** | 33.7 (31.4, 36.3) | 34.4 (32.4, 36.3) | 32.6 (30.3, 36.1) | <0.001 |
| **Arm circumference (cm)** | 28.4 (26.1, 30.7) | 28.5 (26.7, 30.5) | 28.2 (25.1, 31.1) | 0.079 |
| **Handgrip strength (kg)** | 20 (16, 26) | 21 (17, 27) | 20 (15, 25) | 0.011 |
| **Gait speed (s)** | 5.6 (4.5, 7.9) | 5.4 (4.2, 6.9) | 6.4 (4.7, 10.0) | <0.001 |
| **Timed up & go (s)** | 13.7 (10.8, 16.4) | 13.0 (10.4, 15.6) | 14.3 (11.0, 17.3) | 0.084 |
| **Myocardial infarction (%)** | 63 (8.6%) | 35 (8.8%) | 28 (8.4%) | 0.9 |
| **Stroke (%)** | 22 (3.0%) | 13 (3.3%) | 9 (2.7%) | 0.7 |
| **Diabetes (%)** | 157 (21%) | 70 (18%) | 87 (26%) | 0.005 |
| **Hypertension (%)** | 412 (56%) | 215 (54%) | 197 (59%) | 0.2 |
| **Cancer (%)** | 42 (5.7%) | 24 (6.0%) | 18 (5.4%) | 0.7 |
| **Dyslipidemia (%)** | 264 (36%) | 142 (36%) | 122 (36%) | 0.8 |
| **Number of comorbidities** |  |  |  | 0.2 |
| 0 | 185 (25%) | 108 (27%) | 77 (23%) |  |
| 1 | 266 (36%) | 144 (36%) | 122 (36%) |  |
| 2 | 181 (25%) | 101 (25%) | 80 (24%) |  |
| $\geq 3$ | 103 (14%) | 47 (12%) | 56 (17%) | |
| **Death at follow-up** | 61 (10%) | 28 (8.9%) | 33 (12%) | 0.2 |
| Unknown | 149 | 85 | 64 |  |
| **Frailty status at follow-up** |  |  |  | 0.017 |
| Non-fragile | 81 (31%) | 49 (37%) | 32 (24%) |  |
| Pre-frail | 138 (52%) | 67 (51%) | 71 (54%) |  |
| Frail | 44 (17%) | 15 (11%) | 29 (22%) |  |
| Unknown | 472 | 269 | 203 |  |

<sup>1</sup>n (%); Median (IQR), <sup>2</sup>Pearson's Chi-squared test; Wilcoxon rank sum test

### Biological age acceleration and frailty scores

We observed an association between AnthropoAgeAccel and the frailty index (ρ=0.12, 95%CI 0.05-0.19), and between AnthropoAgeAccel and the number of components in the frailty phenotype (ρ=0.17, 95%CI 0.10-0.24, **Figure 2**). We also identified an increase in the frailty index (β=0.0053, 95%CI 0.0027-0.0079) for every 1-year increase in BA acceleration as measured by AnthropoAgeAccel (**Figure 3A**). Additionally, individuals with accelerated aging had higher frailty indices, and we observed an increase in AnthropoAgeAccel with higher number of components of the Fried Frailty Scale, irrespective of sex (**Figures 3B-C**). AnthropoAgeAccel values were also higher in individuals who presented individual frailty phenotype components, except for unintentional weight loss (**Figures 3D-H**).

**Figure 2.**
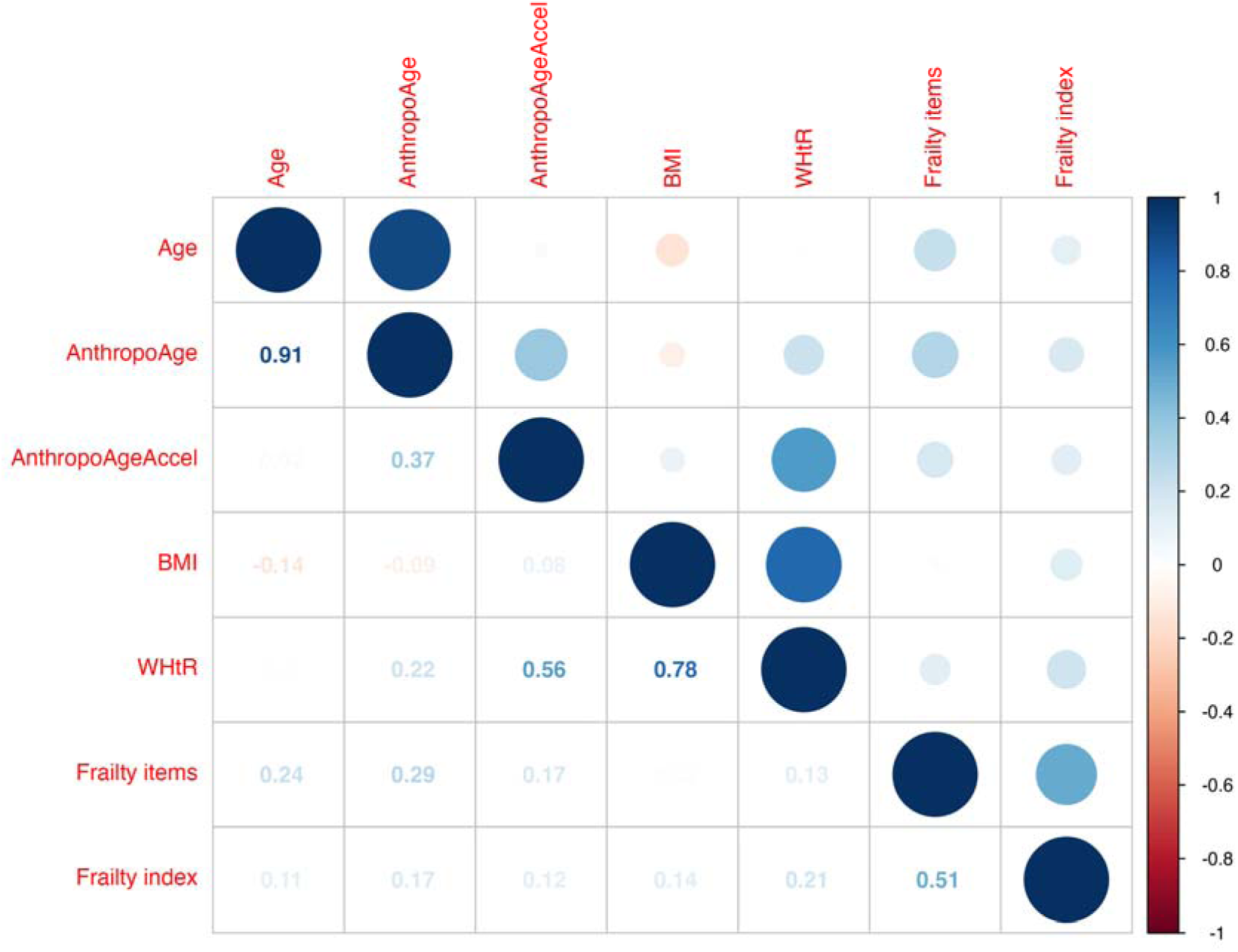
Correlation plot displaying the strength of the linear association of chronological age, AnthropoAge, AnthropoAgeAccel, anthropometric and frailty indices in 735 community-dwelling older adults from the Coyoacán Cohort. ***Abbreviations:*** *BMI, Body-mass index; WHtR, Waist-to-height ratio*

**Figure 3.**
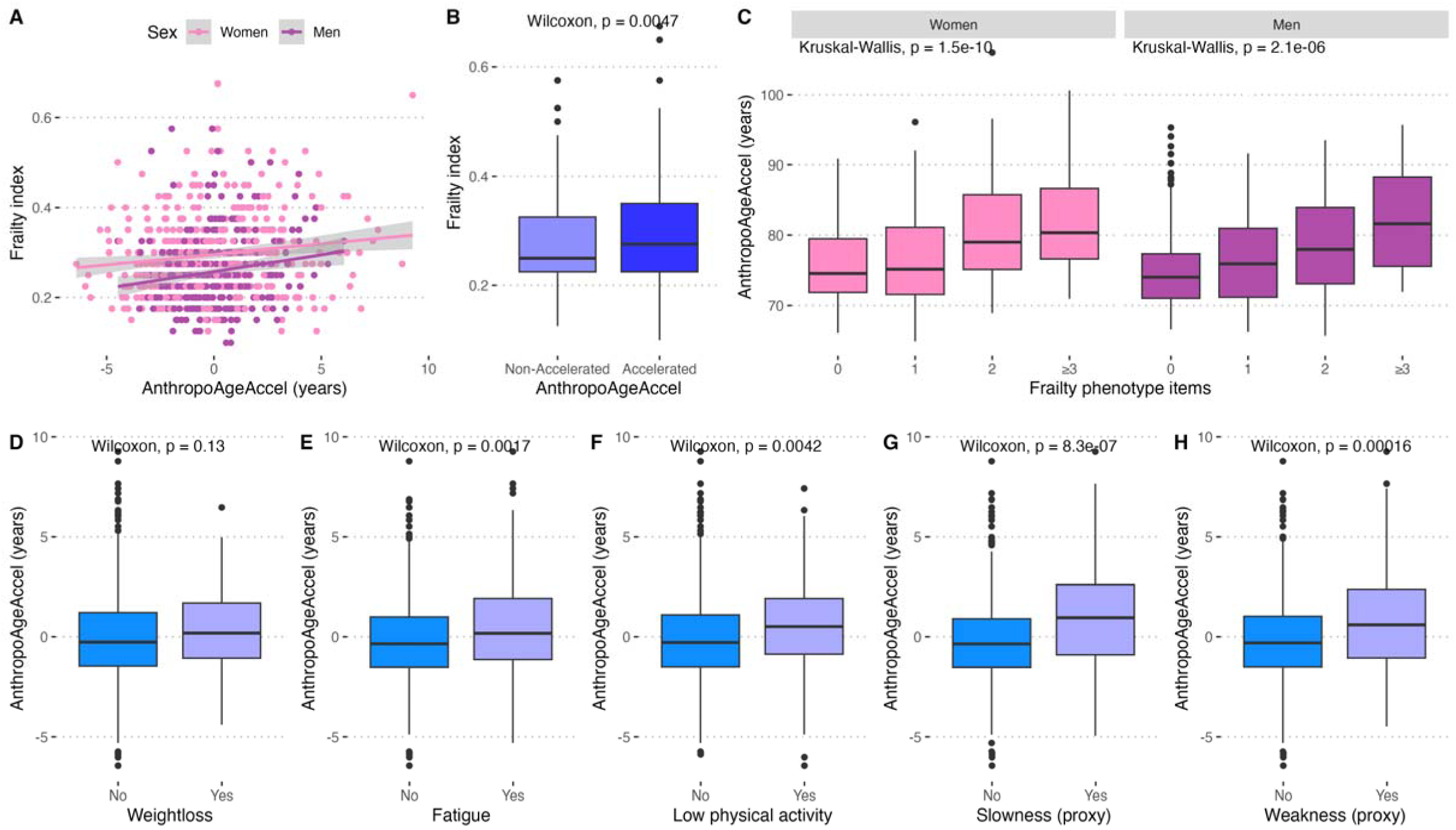
Association between AnthropoAgeAccel with frailty scores in 735 community-dwelling older adults, including the frailty index (**A**), the distribution of frailty indices in individuals with and without accelerated aging, defined as AnthropoAgeAccel values ≥0 years (**B**), as well as the distribution of AnthropoAgeAccel according to number of frailty phenotype items (**C**). The figure also shows comparisons of AnthropoAgeAccel values across individual components of the frailty phenotype (**D-H**).

### Frailty phenotype and accelerated biological aging

Compared to non-frail, frail older adults had an adjusted AnthropoAgeAccel 1.71 years higher (95%CI 1.16, 2.25), without significant differences between pre-frail and frail older adults (β= −0.25 years 95%CI −0.10, 0.60). We estimate that a 1-year increase in AnthropoAgeAccel was associated with ∼16% higher odds (OR 1.16, 95%CI 1.09-1.25), and AnthropoAgeAccel ≥0 with ∼74% higher odds of having a more severe frailty phenotype (OR 1.74, 95%CI 1.31-2.32) at baseline.

### Phenotypes of accelerated biological aging and frailty

At baseline 145/366 (39.6%) non-frail older adults had accelerated aging, and this increased to 133/284 (46.8%) for pre-frail and to 57/85 (67.1%) for frail older adults (**Figure 4A**). When exploring the combined influence of frailty and accelerated aging we observed that in non-frail female participants, those with accelerated aging displayed higher BMI, WHtR, and WHR, they also presented higher arm circumference despite having lower handgrip strength and gait speed, indicating increased adiposity and lower physical function (**Figure 4B**). Conversely, for non-frail male participants, those with accelerated aging had higher WHtR, WHR, similar BMI values and lower calf and arm circumference, along with lower handgrip strength and gait speed, indicating a phenotype of abdominal adiposity, decreased appendicular lean mass, and decreased physical function (**Figure 4C**). Amongst frail and pre-frail participants this sexual dimorphism was absent, and participants with pre-frailty/frailty and accelerated aging all displayed body measures characterized by increased abdominal adiposity, decreased appendicular lean mass, and impaired physical function, irrespective of sex (**Figures 4D-E**).

**Figure 4.**
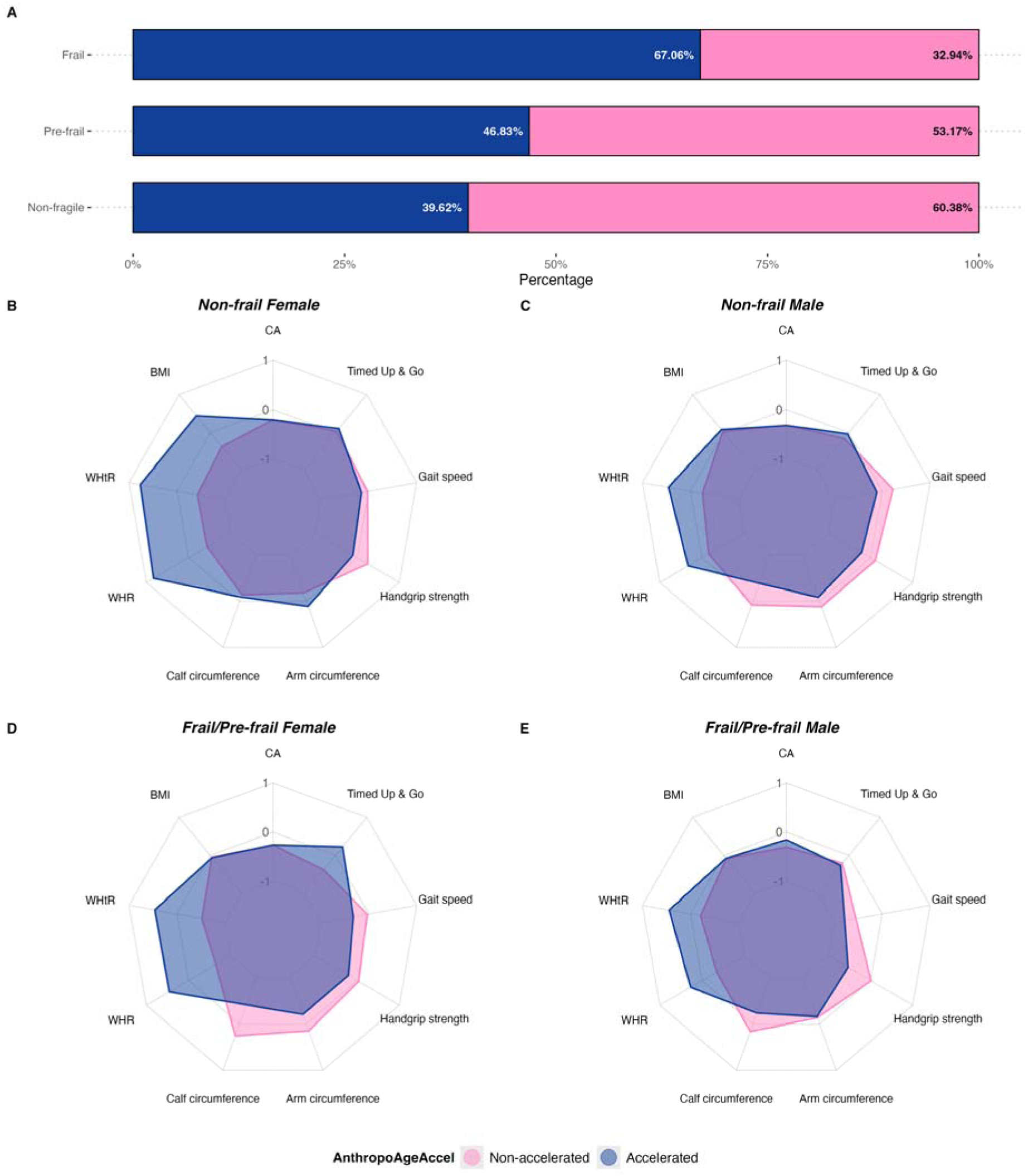
Distribution of accelerated aging defined as AnthropoAgeAccel values ≥0 years in participants according to modified frailty categories at baseline in 735 community-dwelling older adults (**A**). The figure also shows spider plots comparing anthropometry and physical function for participants with and without accelerated aging according to frailty phenotypes and sex (**B-E**). ***Abbreviations:*** *CA, Chronological Age; BMI, Body-mass index; WHtR, Waist-to-height ratio; WHR Waist-to-hip ratio*.

### Influence of accelerated aging on progression to frailty

We analyzed 256 participants with complete 3-year follow-up. Amongst them 134 were non-frail (52.3%), 96 were pre-fail (37.5%), and 26 were frail (10.2%). Amongst non-frail participants 63/134 had accelerated aging at baseline (47.0%), which was lower compared to 47/96 pre-frail (49.0%), and 17/26 frail participants (65.4%, **Figure 5A**). We observed a significant number of transitions across frailty categories over time: participants with frailty increased to 42 (162% increase), pre-frail participants increased to 136 (142% increase), and non-frail participants decreased to 78 (42% decrease, **Figure 5B-C**). A 1-year increase in AnthropoAgeAccel values at baseline predicted ∼19% higher odds of progression from non-frailty to pre-frailty and pre-frailty to frailty at subsequent follow-ups (OR 1.19, 95%CI 1.08-1.31); similarly, participants with accelerated aging at baseline (AnthropoAgeAccel ≥0 years), displayed ∼74% higher adjusted odds of progression to more severe frailty phenotypes at follow-up (OR 1.74, 95%CI 1.11-2.75).

**Figure 5.**
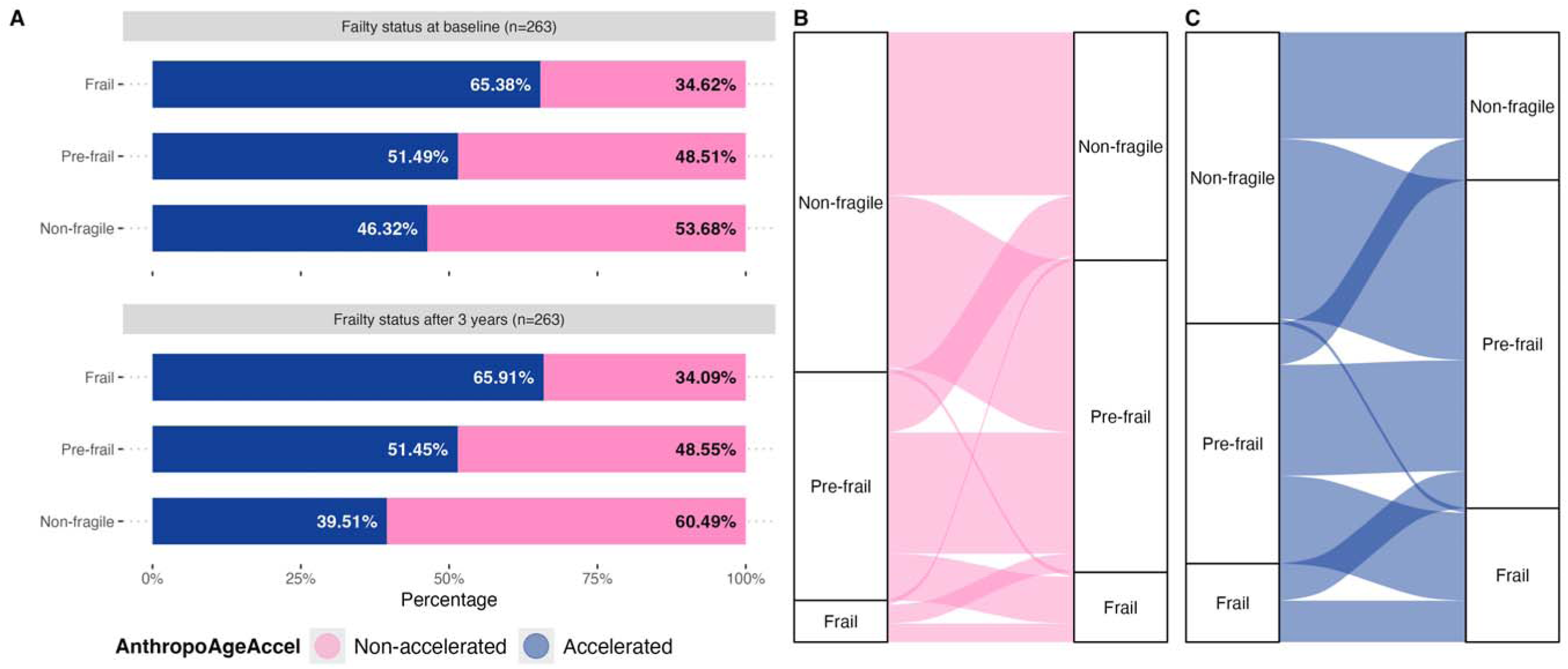
Association between AnthropoAgeAccel with frailty scores including the frailty index (**A**), the distribution of frailty indices in individuals with and without accelerated aging, defined as AnthropoAgeAccel values ≥0 years (**B**), as well as the distribution of AnthropoAgeAccel according to number of frailty phenotype items (**C**).

## DISCUSSION

In this sample of community-dwelling older adults from the Coyoacán Cohort, we characterized accelerated aging as a distinct entity from frailty. Older adults with accelerated aging have higher prevalence of pre-frailty and frailty, as well as lower handgrip strength and gait speed, likely indicative of lower physical function. We also found that older pre-frail or frail adults with accelerated aging had a phenotype indicative of accumulation of visceral adiposity, decreased appendicular lean mass, and decreased physical function, irrespective of sex. This finding is in contrast to our previous study^13^, which showed that accelerated aging displayed a sexual dimorphism in body composition, a finding that we were able to replicate only in non-frail older adults. Finally, we also showed that AnthropoAgeAccel and accelerated aging phenotypes at baseline increased the risk of transitioning over time from non-frail/pre-frail to pre-frail/frail phenotypes, suggesting that BA acceleration is an independent risk factor for frailty progression. Overall, our data suggests that frailty and accelerated BA are intersecting but likely separate phenomena in older adults and that their assessment should be explored separately to better ascertain biologically meaningful aging mechanisms^26,27^.

The distinction between biological aging and frailty has been subject to controversy^2,28,29^. Frailty has been viewed as both a clinical syndrome comprised of low grip strength, slow gait speed, weight loss, exhaustion and low physical activity^20^, as well as an age-related accumulation of health deficits that leads to poor health and increased risk of adverse outcomes^3,24^. Recent evidence suggests that the two definitions of frailty, whilst used interchangeably, are likely expressions of distinct phenomena^26^. Whilst the frailty index captures accumulation of deficits and multisystem deterioration with diverse pathophysiological backgrounds, the frailty phenotype has a more unified pathophysiology and can occur in individuals independent of comorbidity and disability^30,31^. When introducing BA into this assessment, complexity increases as consideration of frailty as an index of deficit accumulation along with epigenetic measures of BA have been shown to be jointly predictive of mortality, indicating that the measures capture distinct but complementary age-related phenomena^32^. In agreement with these findings, out study shows that accelerated BA as captured by AnthropoAge identifies age-related changes that are not fully captured by either the frailty index or the frailty phenotype. We also show that AnthropoAge is useful to identify individuals with impaired physical performance and deficit accumulation in participants before they fulfill criteria for pre-frailty or frailty, and that could be explored as a potentially useful marker for exceptional longevity in older adults^12^. Sarcopenia and decreased muscle function often overlap with frailty in older adults^33,34^. Older frail adults display a body composition phenotype characterized by decreased appendicular lean mass and increased adiposity^35^. Previous research has shown sexual dimorphisms in the contribution of body composition for prediction of frailty over time, with visceral and whole-body adiposity being predictive of frailty in women, but not in men^36,37^. Decreased physical activity, immobility and exhaustion may lead to increased visceral adiposity and decreased muscle mass in frail older adults, thus increasing complexity in the relationship between body composition and frailty^38,39^. In our study, we identified that the sexual dimorphisms of accelerated aging captured by AnthropoAge remain present in older non-frail adults but are not observed in pre-frail or frail older adults. This finding likely indicates that pathophysiological and behavioral changes that occur in pre-frail and frail older adults are influenced by accelerated aging, leading to accumulation of visceral adiposity, decreased muscle mass, and decreased physical function in men and women. Even though we are unable to establish directionality in cross-sectional associations and our cohort spans a short follow-up time, our results call for further longitudinal studies to explore whether accelerated aging impacts body composition changes differentially in frail compared to non-frail older adults.

Our study had several strengths. This is the first study to validate the use of AnthropoAge in a sample of community-dwelling older Mexican adults as a proxy of BA. By using two frailty definitions, we were able to capture both the physical as well as the deficit accumulation phenotype. Finally, by using the longitudinal component of the Coyoacán Cohort study we were able to characterize accelerated aging as a risk factor for frailty progression in older adults, thus establishing AnthropoAgeAccel as a potential frailty marker. We also acknowledge some limitations which should be considered to adequately interpret our results. Despite being an approach previously validated in other studies, the modified frailty phenotype implemented in our study does not make use of objective measures such as grip strength or slow gait, as these measures were only available in a selected subsample of participants. Thus, the strength of the observed associations may have been underestimated, along with the number of frail and pre-frail participants. Moreover, the use of anthropometry to assess body composition only allows approximate inferences on the impact of frailty on the sexual dimorphism in body composition related to accelerated aging, with a need for additional studies based on more precise techniques to explore this phenomenon. Finally, in longitudinal analyses we were able to evaluate changes in the frailty phenotype over time, but not in AnthropoAge or other covariates; despite being able to adjust for their effect at baseline, their dynamic influence over time on frailty progression could not be characterized. Further studies are required to prospectively evaluate the influence of accelerated aging in frailty progression and to explore the utility of AnthropoAge as a complementary measure to assess BA in community-dwelling older adults.

### Conclusions

Our results suggest that, despite being intrinsically intertwined, biological and accelerated aging are phenomena largely independent of frailty both as a phenotype and as an accumulation of age-related deficits. Community-dwelling frail older adults display higher BA acceleration compared to pre-fail and non-frail participants, despite similar CA. Sexual dimorphisms in body composition observed in non-frail participants with accelerated aging are lost in frail older adults with accelerated aging, in whom accumulation of visceral adiposity, decreased appendicular lean mass and physical function are more marked than in frail older adults with non-accelerated aging. Finally, we identified accelerated aging as proxied by AnthropoAgeAccel as a risk factor and a potentially useful biomarker for progression in the severity of the frailty phenotype. Our results are useful to understand the complex interplay between BA, deficit accumulation and physical frailty in older adults and highlight the need for prospective studies to understand how they may capture different aging mechanisms.

## ACKNOWLEDGMENTS

CAFM is enrolled at the PECEM Program of the Faculty of Medicine at UNAM. CAFM and DRG are supported by CONACyT. JAS was supported by Grant Number K23DK135798 from the NIH/NIDDK and by the Massachusetts General Hospital Executive Committee and Center for Diversity and Inclusion Physician-Scientist Development Award

## AUTHOR CONTRIBUTIONS

Research idea and study design: CAFM, CGP, LMGR, OYBC; data acquisition: RCCP, LMGR; analysis/interpretation: CAFM, OYBC; statistical analysis: CAFM, OYBC; manuscript drafting: CAFM, DRG, NEAV, MTLT, RCCP, JAS, CGP, LMGR, OYBC; supervision or mentorship: OYBC. Each author contributed important intellectual content during manuscript drafting or revision and accepts accountability for the overall work by ensuring that questions pertaining to the accuracy or integrity of any portion of the work are appropriately investigated and resolved.

## DATA AVAILABILITY

All code and materials are available for reproducibility of results at https://github.com/oyaxbell/anthropoage_frailty/

